# Automated Text Message Outreach to Increase Diabetes Screening: A Pragmatic Randomized Trial

**DOI:** 10.64898/2026.06.22.26356244

**Authors:** Hayoung Jeong, Leeor Hershkovich, Victoria Glunt, Logan White, Karnika Singh, Matthew J. Crowley, Benjamin A. Goldstein, Anastasia-Stefania Alexopoulos, Jessilyn Dunn

## Abstract

**Background:** Despite evidence that early intervention can prevent or delay progression to type 2 diabetes, more than 80% of individuals with prediabetes in the United States remain undiagnosed, underscoring the need for scalable strategies to increase uptake. In this study, we evaluated whether a single text message could increase completion of HbA1c-based diabetes screening in routine clinical practice.

**Methods:** We conducted a pragmatic randomized controlled trial within Duke University Health System (DUHS). Patients aged 35 years or older who met American Diabetes Association 2022 screening criteria, had no previous diagnosis of diabetes, had not undergone HbA1c testing within the preceding 3 years, and had opted to receive text messages from DUHS were randomly assigned to receive either a single text message encouraging guideline-based diabetes screening and discussion with a primary care provider (intervention group; n=55,494) or usual care (control group; n=5,748). The primary outcome was HbA1c test completion within 24 weeks following message delivery (or no message for controls), analyzed using a Cox proportional hazards model stratified by wave. Secondary outcomes included piecewise hazard ratios for early (weeks 1–4), mid (weeks 5–12), and late (weeks 13–24) intervals and the between-group difference in cumulative testing rate.

**Findings:** Text message outreach significantly increased HbA1c test completion over 24 weeks (HR, 1.18 [95% CI, 1.07–1.03]) with the strongest effect in the first four weeks (HR, 1.48 [95% CI, 1.18–1.86]). By the end of the 24-week observation period, cumulative testing reached 9.14% in the messaged group vs 7.83% in controls (between-group difference, 1.31% [95% CI, 0.59–2.07]), corresponding to one additional HbA1c test per 76 messages delivered ($0.51 in messaging costs per additional HbA1c test performed). Rates of prediabetes and diabetes among those screened were similar between groups, indicating no selection bias of higher-risk patients. One additional dysglycemia case was identified per 213 messages sent ($1.43 per case detected).

**Interpretation:** A single automated text message can increase diabetes screening completion at low marginal cost without selectively engaging higher-risk patients. This low-cost, highly scalable approach could be used to narrow the gap between current and recommended diabetes screening.

## Introduction

Type 2 diabetes (T2D) affects over 40 million individuals in the United States (US)^1^ and is associated with cardiovascular disease, nephropathy, retinopathy, and premature mortality^2,3^.T2D is often preceded by prediabetes (PD), an intermediate state in which timely lifestyle interventions such as increased physical activity and dietary modification can prevent, delay, or even reverse disease progression. Yet, more than 25% and 80% of US adults with diabetes and PD, respectively, are unaware of their condition^1,4^.

In response to the rising incidence of T2D in younger populations, the American Diabetes Association (ADA) updated its PD and T2D screening guidelines in 2022, lowering the recommended age for routine screening from 45 to 35 years for all asymptomatic adults without risk factors, such as family history or comorbidities^5–8^. Recent estimates using National Health and Nutrition Examination Survey (NHANES) data suggest that, compared with the 2003 ADA criteria, the updated guideline expands eligibility among US adults by 6 percentage points (77% vs 83% of the US adults) and that fewer individuals with an unrecognized abnormal HbA1c would be missed under the expanded criteria^9,10^. However, only 47% of US adults who were eligible received glucose testing within the preceding three years, with underscreening disproportionately affecting younger adults, Hispanic individuals, and those without a regular source of care^9^.

Scalable, low-cost outreach strategies are needed to engage large, diverse populations of eligible individuals outside of traditional visit-based care. Text message-based outreach has been shown to improve appointment attendance, medication adherence, cancer screening, and health behaviors across diverse settings^11–14^. In addition, text message outreach costs substantially less than phone call reminders, especially when accounting for additional staff time, while achieving similar effectiveness^15,16^.

In this study, we conducted a pragmatic randomized controlled trial evaluating the effectiveness of a single automated text message in increasing diabetes screening rates within a large academic health system.

## Methods

### Study design

We conducted a diabetes awareness campaign within the Duke University Health System (DUHS) as an IRB-exempt quality improvement study (IRB Protocol#: 00112670). We queried our electronic health record (EHR) database using predefined eligibility criteria consistent with ADA screening guidelines to identify patients^6^. Eligibility included age ≥35 years, no known prior diagnosis of T2D or PD recorded in the EHR (ICD codes R73.xx and E11.xx) or the North Carolina Diabetes Registry, and no documented HbA1c test within the preceding three years.

We only included patients who previously provided consent for health-system-based text message communication, and who had ≥1 primary care visit(s) within the prior 24 months (to ensure that they were receiving primary care in DUHS and were likely to have their follow-up HbA1c test done within the health system) (Figure 1). Outreach was deployed in three sequential waves between March 2025 and May 2025 with prospectively randomized control groups. Twenty-two thousand patients were sampled in each wave and randomized in a 10:1 ratio, with 20,000 eligible patients assigned to receive the text outreach and 2,000 serving as contemporaneous controls, for a total of 66,000 patients. The 10:1 allocation ratio was selected pragmatically to maximize the number of patients receiving the intervention. The pooled sample across the three waves (60,000 intervention and 6,000 control patients) provided approximately 80% power to detect a between-group difference of 1.2% (two-sided α = 0.05), based on a testing rate of 11.9% observed in a prior single-arm pilot study (N = 2,976). Randomization was performed using a computer-generated procedure. Messages were deployed through a HIPAA-compliant text messaging platform already integrated within DUHS at a marginal cost of $0.0067 per message. Eligible patients randomized to the messaged group received a single text message: “The ADA recommends that everyone aged 35 and older be screened for diabetes every 3 years. As you prepare for your next visit, please consider discussing diabetes screening with your Primary Care Provider. Learn more at our website [link to study website]” (Supplementary Figure 1).

**Figure 1.**
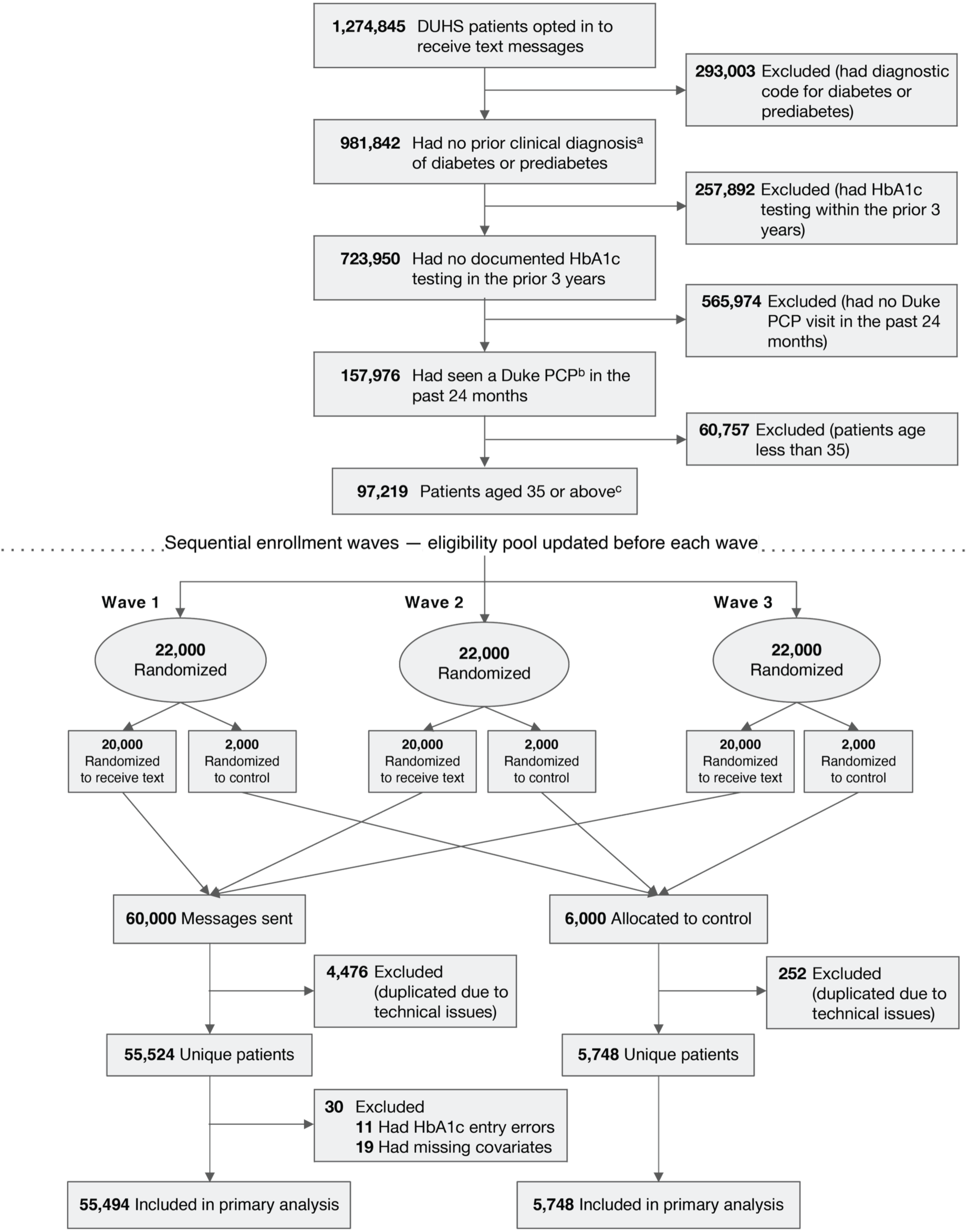
Flow of participants through the study.

### Analytic cohort

All analyses followed an intent-to-treat approach, including patients regardless of message delivery status. We extracted patient age, sex, race, and ethnicity from the EHR. In both arms, we excluded duplicate patient records arising from messaging system artifacts and patients with entry errors in HbA1c values (i.e., non-numeric or HbA1c > 50) or missing all demographic variables to enable subgroup analysis.

### Outcomes

The primary outcome was completion of HbA1c testing within 24 weeks, as recorded in the DUHS EHR system, including tests conducted both within and outside of DUHS that were documented in the EHR. Secondary outcomes included the proportion of patients with newly identified prediabetes and diabetes (i.e., dysglycemia) following testing. Testing outcomes were classified based on the first HbA1c result recorded during the 24-week observation period: Normoglycemia (HbA1c < 5.7%), Prediabetes (HbA1c 5.7%-6.4%), Diabetes (HbA1c ≥ 6.5%), in accordance with ADA criteria^17^.

### Statistical analysis

We assessed whether there were demographic differences between the messaged and control (not messaged) groups using the standardized mean difference (SMD). We estimated the cumulative incidence of HbA1c testing as the complement of the Kaplan-Meier survival estimate (1 − KM) and calculated the group difference at 24 weeks with 95% confidence intervals obtained via bootstrapping. The primary outcome was analyzed as a time-to-event outcome using a Cox proportional hazards model stratified by outreach wave, with follow-up censored at 24 weeks. To examine variation in intervention effects across demographic subgroups, we conducted stratified analyses by sex, race, ethnicity, and age group. We also used Piecewise Cox models to estimate hazard ratios across prespecified early (weeks 1–4), mid (weeks 5–12), and late intervals (weeks 13–24). All analyses were conducted in Python (version 3.11.5) using lifelines package (version 0.30.0)^18^. As a secondary analysis, we examined differences in the diagnostic outcomes among tested individuals between the messaged and control groups using the Chi-square test.

Lastly, we derived the number needed to message (NNM) to prompt one additional screening completion. The NNM was estimated by taking the inverse of the group difference in HbA1c completion rates between the intervention and control groups and multiplying by the marginal cost per message ($0.0067). The NNM to identify one additional case of dysglycemia (PD or T2D) was calculated by dividing the screening NNM by the proportion of screened patients in the intervention arm with an above normal HbA1c result.

## Results

### Study population

The final analytic cohort included 55,494 patients in the messaged group and 5,748 patients in the control group (Figure 1). Among the 60,000 initially sampled patients who received the text message, 4,476 (7.5%) had multiple message attempts due to technical errors, as patients with indeterminate or failed delivery status were returned to the eligibility pool and resampled in subsequent waves. In the control group, 252 patients (4.2%) were duplicated across waves; these individuals were excluded to ensure mutually exclusive exposure groups and a nonoverlapping analytic cohort. Additional exclusions included patients with nonnumeric or implausible HbA1c values (n=11) and missing demographic data (unknown sex or age; n=19). Baseline demographic characteristics were balanced between the text message and control groups, with no SMD greater than 0.10. (Table 1).

**Table 1.**
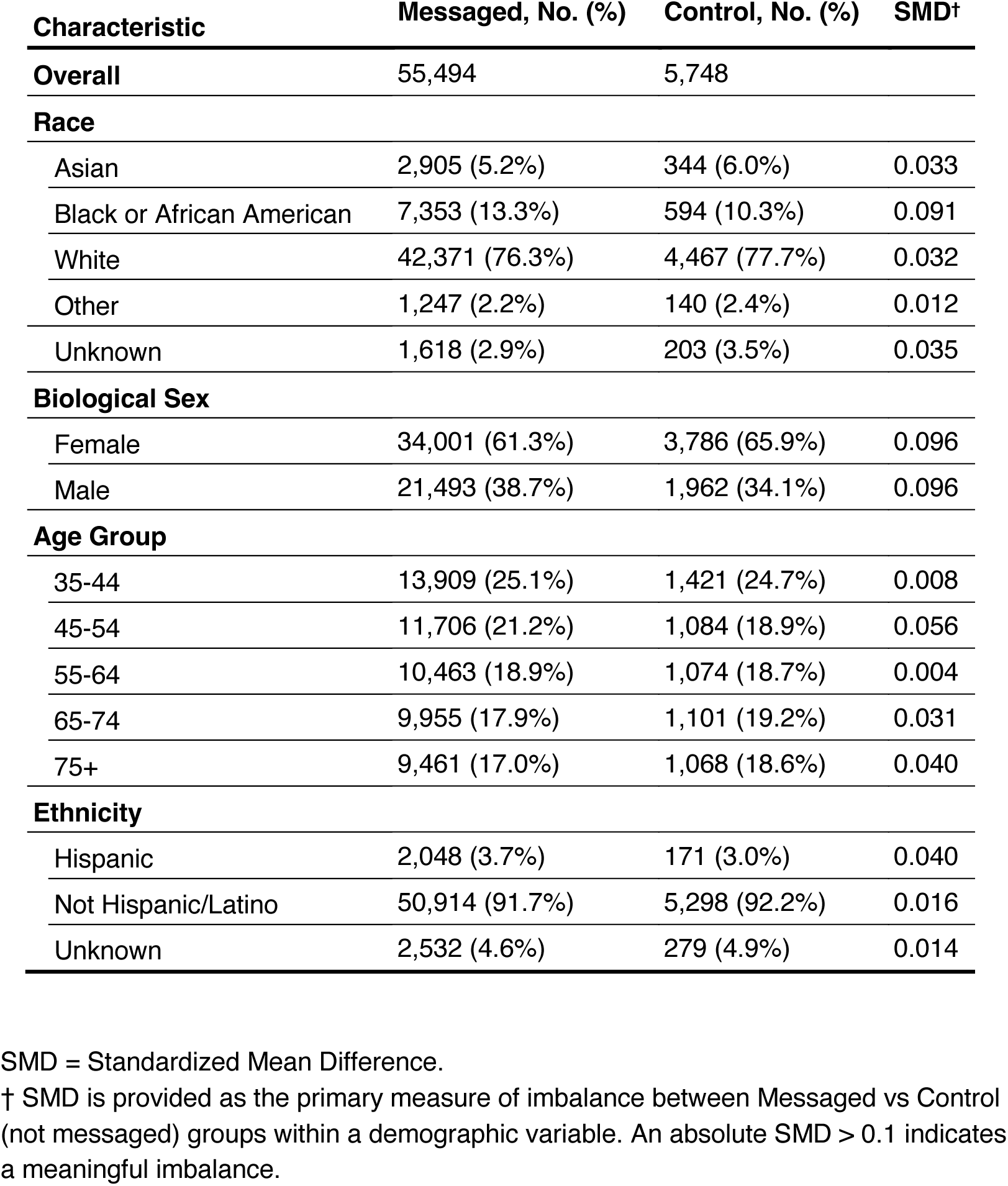
Demographic characteristics of the study population included in the analysis.

### Effectiveness of the SMS Outreach Campaign

At the end of the 24-week observation period, cumulative screening reached 9.14% (95% CI, 8.91-9.39) in the messaged group compared with 7.83% (95% CI, 7.16-8.55) in the control group (HR, 1.18 [95% CI, 1.07–1.30]; between-group difference, 1.31% [95% CI, 0.59-2.07]) (Figure 2). Testing rates were 48% higher in the messaged group than the control group in the first four weeks (HR, 1.48 [95% CI, 1.18–1.86]). This effect attenuated in the mid period (HR, 1.20 [95% CI, 1.01–1.41]) and was no statistically significant difference in the rate of new screening between the messaged and control groups in the late period (HR, 1.10 [95% CI, 0.96–1.27]) (Figure 3). Effects were directionally consistent across all demographic subgroups (Supplementary Figure 2).

**Figure 2.**
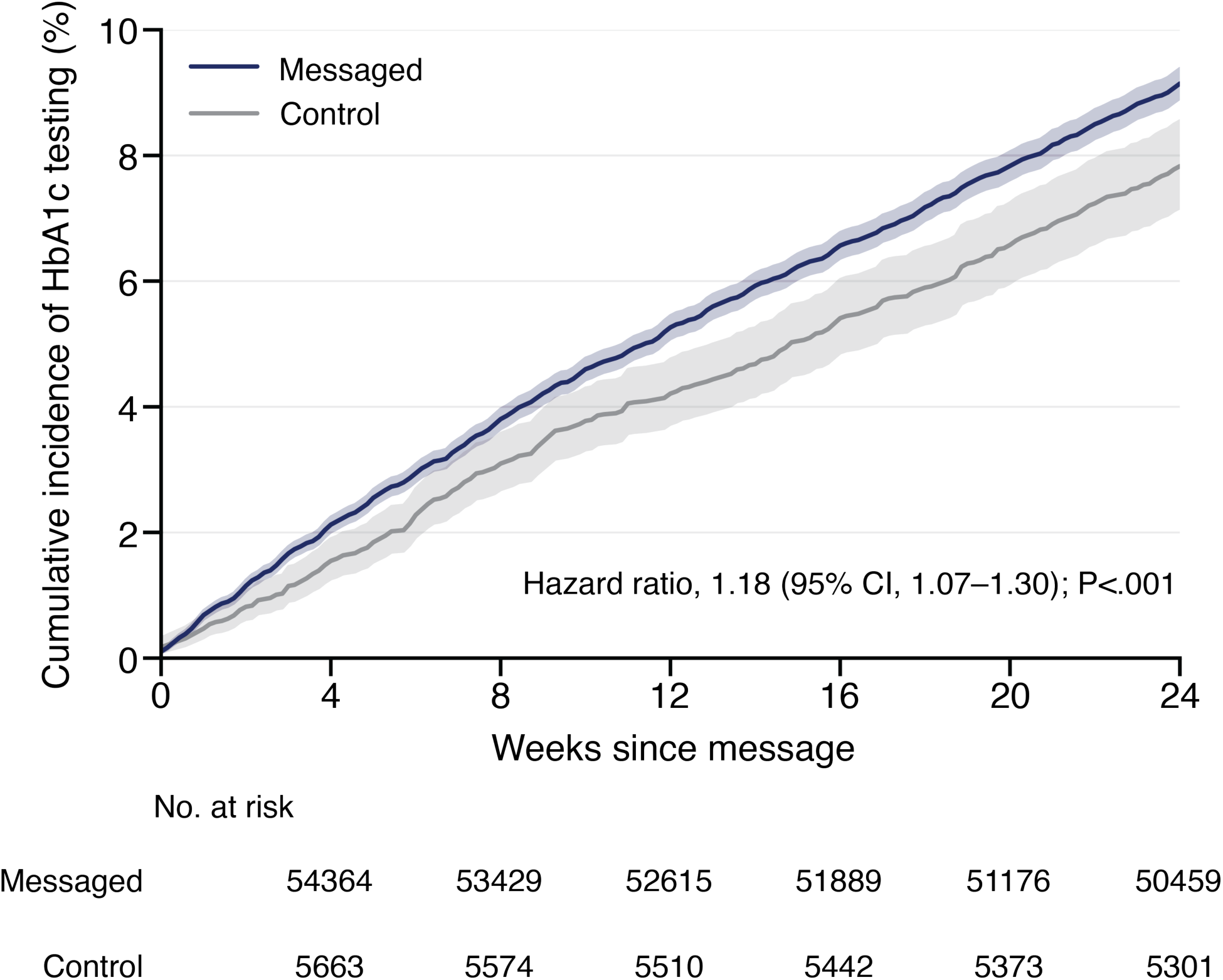
Cumulative incidence curve of HbA1c testing uptake following the text message outreach. Cumulative incidence of HbA1c testing over 24 weeks among patients who received a text message encouraging diabetes screening (Messaged, n=55,494) and those without any intervention (Control, n=5,748) was derived as the complement of the Kaplan-Meier survival estimate (1 – KM estimate). Shaded areas indicate 95% confidence intervals (CI). Hazard ratio and P value are from a Cox proportional hazards regression model stratified by messaging wave.

**Figure 3.**
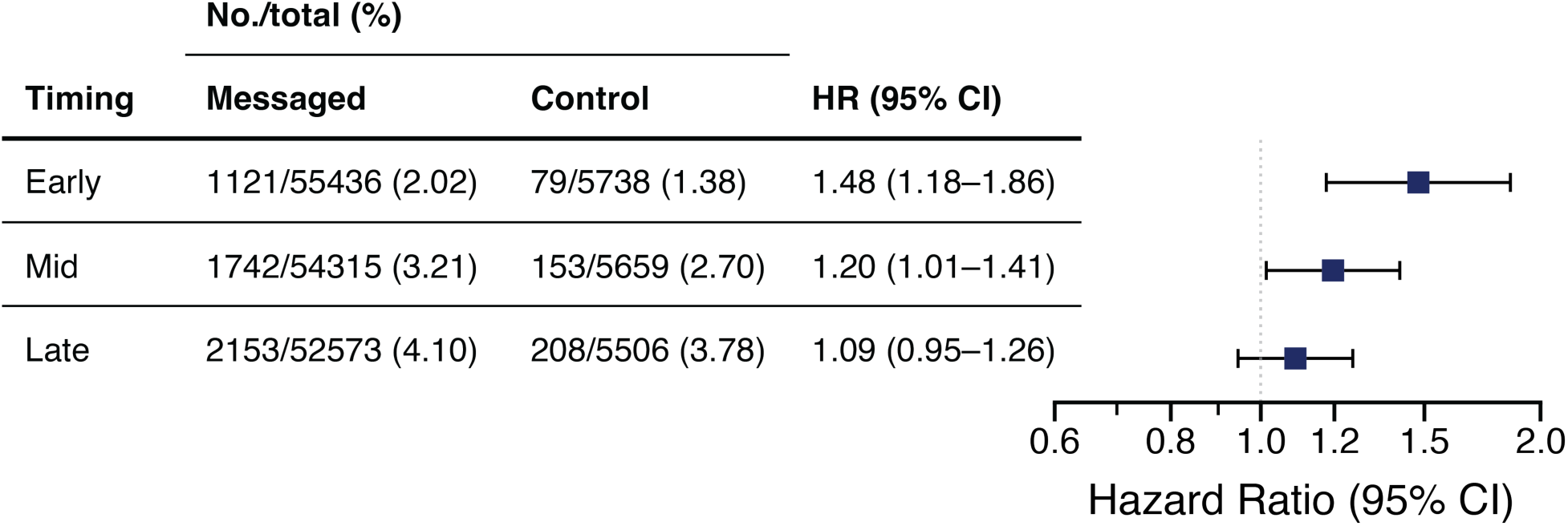
Time-varying association of text message outreach with HbA1c testing completion. Hazard ratios (HR) and 95% confidence intervals (CI) for HbA1c testing are shown across prespecified time intervals following outreach: early (weeks 1–4, excluding the day of message), mid (weeks 5–12), and late (weeks 13–24). HRs were estimated from a Cox proportional hazards model stratified by outreach wave. Event counts for the messaged and control groups are shown for each interval.

### Newly Identified Prediabetes and Type 2 Diabetes

Among individuals who completed HbA1c testing (messaged, n = 5,074; control, n = 450), the proportion of test results indicative of PD or T2D was similar between the messaged and control groups (χ² test, P = 0.865), with comparable distributions of prediabetes (33.6% vs 32.9%) and diabetes (2.1% vs 2.4%) (Figure 4). Among adults aged 35–44 who became newly eligible for PD/T2D screening under the updated ADA guidelines^6,17^ (n=1,281 messaged, n=118 control), nearly one in five (19.0%) had above normal HbA1c result (18.1% PD or 0.9% T2D) (Supplementary Table 1).

**Figure 4.**
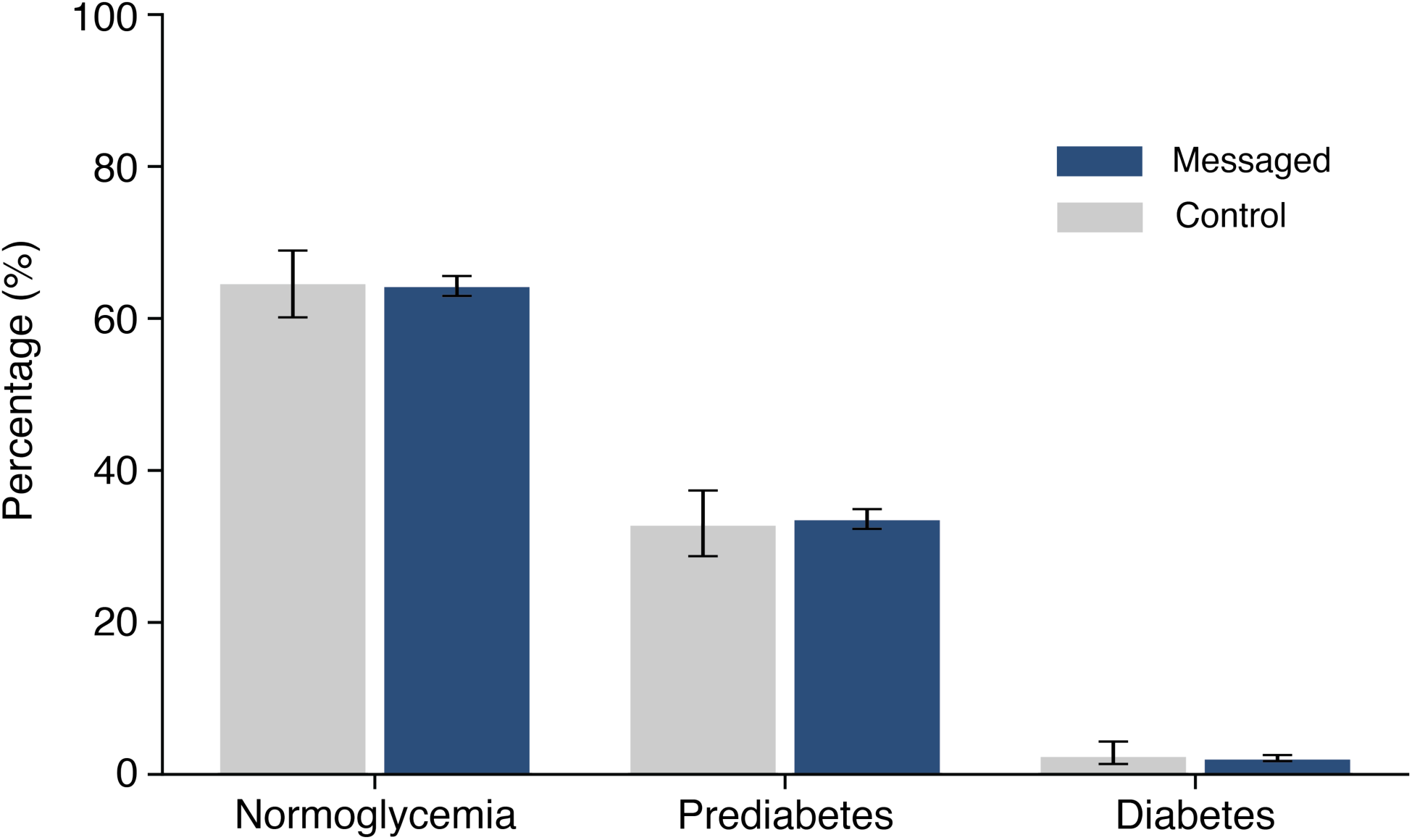
HbA1c testing outcomes among those who tested during the 24-week observation period. Error bars indicate 95% confidence intervals calculated using the Wilson method (control n = 450; messaged n=5,074).

The messaged group’s increased testing led to a higher case detection of newly identified prediabetes (3.10% vs 2.57%) and normoglycemic results (5.93% vs 5.06%), whereas rates of newly identified diabetes were low and did not differ between groups (0.19% in both groups) (Supplementary Table 2).

### Number Needed to Message and Cost of Outreach

Overall, the 1.31% between-group difference in testing rates translates to approximately one additional completed HbA1c test for every 76 patients who each received a single text message. Given an approximately 36% rate of abnormal HbA1c results in the messaged group, one additional case of dysglycemia was detected for every 213 messages sent. The total marginal cost of the text outreach was approximately $400 for delivering 60,000 messages. At less than $0.01 per-message cost, the outreach corresponded to $0.51 per additional completed HbA1c test. The cost per additional case of dysglycemia identified was $1.43.

## Discussion

In this pragmatic randomized controlled trial, a single text message significantly increased the completion of HbA1c testing among at-risk patients who had not undergone screening for diabetes. Over 24 weeks, messaged patients completed HbA1c testing at an 18% higher rate than controls (HR 1.18), with the effect most pronounced in the first four weeks (HR 1.48). This resulted in a 1.31% between-group difference in cumulative screening. Text outreach did not appear to selectively reach or motivate higher-risk patients, with approximately 36% of screened patients in the messaged group having an abnormal HbA1c test, similar to the rate in the control group. Although the absolute difference in screening completion was modest from this single text message intervention, the scalability of text-based outreach suggests potential for meaningful population-level impact.

The cost profile of this intervention strengthens the case for adoption. At less than $0.01 per message, every 76 messages sent prompted one additional completed HbA1c test ($0.51 per additional screening), and every 213 messages identified one additional case of dysglycemia ($1.43 per case detected). These figures compare favorably with more resource-intensive outreach modalities, such as phone calls, which may require substantial staff time and also be more likely to be missed. Given that many healthcare systems already employ HIPAA-compliant text messaging (e.g., for appointment reminders), few to no additional resources would be required to implement such SMS outreach for screening.

Our study provides a benchmark for large-scale digital outreach campaigns to promote diabetes screening. Within diabetes care, text-based reminders have demonstrated meaningful improvements in appointment attendance^11,19^, medication adherence^13^, and postpartum glucose tolerance testing completion among women with gestational diabetes^12^. While evidence of text interventions has also increased uptake of preventive cancer screenings^14,20^, the utility of automated text outreach for promoting routine diabetes screening in at-risk patients had not been established; to our knowledge, this is the first randomized controlled trial to demonstrate its effectiveness in this context. In primary care systems, where clinician-led outreach is often resource-intensive, our findings support automated text as a scalable tool for identifying undiagnosed PD or T2D at low marginal cost, providing an otherwise missing window of opportunity for care.

The responsiveness we observed in our text message campaign suggests a degree of digital engagement in this population that could extend to mobile-based lifestyle interventions. There is growing recognition of the importance of virtual lifestyle interventions and education for PD/T2D as a way to personalize care approaches and better meet patients where they are. For instance, the CDC’s National Diabetes Prevention Program (DPP) is geared towards preventing T2D in individuals at high risk (e.g., PD), and the DPP has shifted from being delivered solely as an in-person program to increasingly offering virtual programs. Evidence suggests potentially greater participation in virtual as compared to in-person DPP, with similar weight loss at one year^21^. The Diabetes Prevention Alliance is a recent CDC/ADA initiative designed to more effectively connect individuals with PD to eligible DPP offerings, the majority of which are delivered virtually^22^. In addition, for individuals diagnosed with T2D, the 2026 ADA guidelines now acknowledge digital self-management tools and coaching as appropriate methods of support^23^.

In this context, text-based outreach can function not only as a screening prompt but also as a gateway into a broader digital care pathway for patients newly identified as having PD or T2D.

While our approach relied on population-wide text messaging, integrating clinical risk factors beyond the age criteria (e.g., family medical history or BMI), prior utilization patterns, or patient-generated data sources such as wearable devices could enable targeted outreach to individuals at the highest risk, potentially improving both screening uptake and specificity (i.e., positivity rates)^24,25^. Future studies should also evaluate adaptive, personalized messaging strategies and deployment across diverse health systems to optimize the reach and impact of digital screening programs. The text outreach employed a standardized, non-personalized, one-time message that did not account for the patient’s digital literacy or behavioral motivation, nor did it include any follow-up messages if no action was taken. Future systems could incorporate adaptive designs that modify content and cadence based on individual response patterns or previous clinical encounters^26^. In addition, although not currently used for definitive diagnosis, remote testing strategies (e.g., HbA1c test kits by mail) could complement digital outreach by reducing logistical barriers to initial screening, particularly in underserved or rural populations with limited access to in-person care^27–30^.

The scalability and low marginal cost of text message-based outreach make it particularly well-suited for population-wide screening initiatives. The efficiency of the text outreach—measured by the number of messages required to identify a new case—suggests meaningful returns when deployed at scale. In settings facing increasing preventive care demands, automated digital outreach may complement clinician-led efforts and help close persistent gaps in screening for a condition that remains substantially underdiagnosed.

We did not capture whether patients requested HbA1c tests but did not receive them—clinicians may have declined patient requests to order tests based on clinical judgement or concerns about insurance coverage, potentially attenuating observed screening uptake independent of patient action. Although the exploratory analyses suggest directionally consistent results across demographic strata, the study was not powered to detect differential treatment effects within subgroups, and estimates for smaller populations (e.g., Hispanic or Asian patients) were imprecise, with wide confidence intervals that frequently included the null. As a result, conclusions about heterogeneity of treatment effect by race, ethnicity, or other characteristics should be interpreted with caution and confirmed in adequately powered studies. In addition, some patients may have completed their HbA1c testing outside of DUHS, and as a result, their test may have not been documented in the DUHS EHR. We also limited eligibility based solely on HbA1c screening as opposed to other diabetes screening methods (like plasma glucose, because fasting status is inconsistently captured in EHR data for glucose testing). As a result, some patients who were reached out to may have had blood glucose tests other than HbA1c performed either before or after receiving the text message, neither of which would have been captured here, indicating that screening rates may actually be higher than what was measured through our methods. The study was conducted within a single academic health system located in the Southeastern US, which may limit generalizability to other settings with different patient populations. Study enrollment was restricted to patients who had previously consented to receive text messages from DUHS. Patients who opt in may be more motivated to engage with their healthcare, potentially overestimating the effectiveness of this intervention in broader populations.

In this pragmatic randomized controlled trial, a single text message significantly increased diabetes screening uptake among patients eligible under the recently expanded ADA screening criteria. These findings support automated SMS outreach as a highly scalable, low-cost, and low-lift strategy for translating evidence-based screening recommendations into clinical action.

### Contributors

HJ, MJC, BAG, AA, and JD conceived and designed the study. HJ, LH, VG, LW, MJC, BAG, AA, and JD contributed to data acquisition, analysis, or interpretation. HJ, AA, and JD drafted the manuscript. MJC, BAG, AA, and JD critically revised the manuscript for important intellectual content. HJ performed the statistical analyses under BAG, and JD supervision. VG, LW, and KS provided administrative, technical, or material support. JD supervised the study. JD had full access to all the data in the study and takes responsibility for the integrity of the data and the accuracy of the data analysis. All authors contributed to interpretation of the findings, reviewed the manuscript, and approved the final version.

### Data Sharing

The individual-level participant data underlying this study will not be made publicly available because of institutional and privacy restrictions related to protected health information.

Aggregate study results are reported in the manuscript and supplementary materials.

## Data Availability

The individual-level participant data underlying this study will not be made publicly available because of institutional and privacy restrictions related to protected health information. Aggregate study results are reported in the manuscript and supplementary materials.

## Declaration of Interests

JD sits on the Google Consumer Health Advisory Board and is a consultant to Samsung Research America and Jones Day. Other authors declare no competing interests.

## Acknowledgements

This study was supported by the NIH/NIDDK R01 DK133531 award. MJC acknowledges funding from the National Institutes of Health (1R01NR019594), Veterans Affairs Quality Enhancement Research Initiative (VA QUE 25-017), and the Veterans Affairs Office of Rural Health (NOMAD 03480). HJ’s effort was supported by NHLBI F31HL179990.

## Notes

### Author Declarations

The study protocol was reviewed by the Duke IRB and determined to be an IRB-exempt quality improvement study (IRB Protocol#: 00112670).

## References

1 CDC. A U.S. Report Card. Diabetes. 2026; published online March 16. https://www.cdc.gov/diabetes/communication-resources/diabetes-statistics.html (accessed May 10, 2026).

2 Tomic D, Shaw JE, Magliano DJ. The burden and risks of emerging complications of diabetes mellitus. Nature Reviews Endocrinology 2022; 18: 525–39.

3 Zheng Y, Ley SH, Hu FB. Global aetiology and epidemiology of type 2 diabetes mellitus and its complications. Nature Reviews Endocrinology 2018; 14: 88–98.

4 CDC. Prediabetes: Could It Be You? Diabetes. 2026; published online Feb 17. https://www.cdc.gov/diabetes/communication-resources/prediabetes-statistics.html (accessed May 10, 2026).

5 CDC. Additional 12 Million U.S. Adults Eligible for Diabetes Screening. Diabetes. 2026; published online Jan 26. https://www.cdc.gov/diabetes/data-research/research/diabetes-screening-eligible.html (accessed May 10, 2026).

6 American Diabetes Association Professional Practice Committee. 2. Classification and Diagnosis of Diabetes: Standards of Medical Care in Diabetes—2022. Diabetes Care 2021; 45: S17–38.

7 Chung S, Azar KMJ, Baek M, Lauderdale DS, Palaniappan LP. Reconsidering the Age Thresholds for Type II Diabetes Screening in the U.S. American Journal of Preventive Medicine 2014; 47: 375–81.

8 US Preventive Services Task Force. Screening for Prediabetes and Type 2 Diabetes: US Preventive Services Task Force Recommendation Statement. JAMA 2021; 326: 736–43.

9 Ali MK, Imperatore G, Benoit SR, et al. Impact of changes in diabetes screening guidelines on testing eligibility and potential yield among adults without diagnosed diabetes in the United States. Diabetes Res Clin Pract 2023; 197: 110572.

10 Fang M, Wang D, Echouffo-Tcheugui JB, Selvin E. Prediabetes and Diabetes Screening Eligibility and Detection in US Adults After Changes to US Preventive Services Task Force and American Diabetes Association Recommendations. JAMA 2022; 327: 1924–5.

11 Liew S-M, Tong SF, Mun Lee VK, Ng CJ, Leong KC, Teng CL. Text messaging reminders to reduce non-attendance in chronic disease follow-up: a clinical trial. Br J Gen Pract 2009; 59: 916–20.

12 Milluzzo A, Manuella L, Frittitta L, Sciacca L. Efficacy of a phone reminder to improve adherence to post-partum glucose tolerance testing after gestational diabetes and clinical predictors of post-partum follow-up compliance. https://www.diabetesresearchclinicalpractice.com/article/S0168-8227(24)00563-1/fulltext (accessed May 10, 2026).

13 Belete AM, Gemeda BN, Akalu TY, Aynalem YA, Shiferaw WS. What is the effect of mobile phone text message reminders on medication adherence among adult type 2 diabetes mellitus patients: a systematic review and meta-analysis of randomized controlled trials. BMC Endocr Disord 2023; 23: 18.

14 Schwebel FJ, Larimer ME. Using text message reminders in health care services: A narrative literature review. Internet Interventions 2018; 13: 82–104.

15 Bracken K, Keech A, Hague W, et al. Telephone call reminders did not increase screening uptake more than SMS reminders: a recruitment study within a trial. Journal of Clinical Epidemiology 2019; 112: 45–52.

16 Gurol-Urganci I, De Jongh T, Vodopivec-Jamsek V, Atun R, Car J. Mobile phone messaging reminders for attendance at healthcare appointments. Cochrane Database of Systematic Reviews 2013; 2013. DOI:10.1002/14651858.CD007458.pub3.

17 American Diabetes Association. Standards of Medical Care in Diabetes—2022 Abridged for Primary Care Providers. Clin Diabetes 2022; 40: 10–38.

18 Davidson-Pilon C. lifelines, survival analysis in Python. 2024; published online Oct 29. DOI:10.5281/zenodo.14007206.

19 Wong ZKH, Chai XY, Aqilah MN, et al. SMS Reminders for Increasing Follow-up Adherence among Diabetic Patients-an Innovative Strategy. Prim Health Care 2019; 9.

20 Uy C, Lopez J, Trinh-Shevrin C, Kwon SC, Sherman SE, Liang PS. Text Messaging Interventions on Cancer Screening Rates: A Systematic Review. J Med Internet Res 2017; 19: e296.

21 Moin T, Damschroder LJ, AuYoung M, et al. Results From a Trial of an Online Diabetes Prevention Program Intervention. Am J Prev Med 2018; 55: 583–91.

22 Learn About the National Diabetes Prevention Program | American Diabetes Association. https://diabetes.org/about-diabetes/diabetes-prevention/dpp (accessed May 13, 2026).

23 American Diabetes Association. 3. Comprehensive Medical Evaluation and Assessment of Comorbidities. Diabetes Care 2017; 40: S25–32.

24 Sahin C, Courtney KL, Naylor P, E Rhodes R. Tailored mobile text messaging interventions targeting type 2 diabetes self-management: A systematic review and a meta-analysis. Digit Health 2019; 5: 2055207619845279.

25 Dong J, Li JX, Aung N, Smith C, Anderson TS. Comparison of Generic Versus Personalized Text Messages for Diabetes Laboratory Monitoring: a Randomized Quality Improvement Study. J Gen Intern Med 2023; 38: 2001–2.

26 Ramirez M, Wu S, Beale E. Designing a Text Messaging Intervention to Improve Physical Activity Behavior Among Low-Income Latino Patients With Diabetes: A Discrete-Choice Experiment, Los Angeles, 2014–2015. Prev Chronic Dis 2016; 13: 160035.

27 AlQassab O, Kanthajan T, Pandey M, et al. Evaluating the Impact of Telemedicine on Diabetes Management in Rural Communities: A Systematic Review. Cureus 2024; published online July 19. DOI:10.7759/cureus.64928.

28 Delaronde S. Barriers to A1C Testing Among a Managed Care Population. Diabetes Educ 2005; 31: 235–9.

29 Chang A, Frank J, Knaebel J, Fullam J, Pardo S, Simmons DA. Evaluation of an Over-the-Counter Glycated Hemoglobin (A1C) Test Kit. J Diabetes Sci Technol 2010; 4: 1495–503.

30 Singh K, Esteve L, North P, et al. Unmasking Hidden Dysglycemia: A Mobile OGTT Approach Using Continuous Glucose Monitors. medRxiv 2025; : 2025.12.14.25341915.

