## Supplementary for "Automated Text Message Outreach to Increase Diabetes Screening: A Pragmatic Randomized Trial"


Supplementary Table 1 2

Supplementary Table 2 3

Supplementary Figure 1 4

Supplementary Figure 2 5

**Supplementary Table 1. Distribution of glycemic categories among individuals who completed HbA1c testing within 24 weeks, stratified by age group.**

| | | | Number of patients (Proportion, %) | | |
|---|---|---|---|---|---|
| **Age Group** | **Group** | **Total, n** | **Normoglycemia** | **Prediabetes** | **Diabetes** |
| 35-44 | Control | 188 | 93 (78.8) | 25 (21.2) | 0 |
| | Messaged | 1281 | 1040 (81.2) | 228 (17.8) | 13 (1.0) |
| | Total | 1469 | 1133 (81.0) | 253 (18.1) | 13 (0.9) |
| 45-54 | Control | 80 | 54 (67.5) | 24 (30.0) | 2 (2.5) |
| | Messaged | 1034 | 730 (70.6) | 287 (27.8) | 17 (1.6) |
| | Total | 1114 | 784 (70.4) | 311 (27.9) | 19 (1.7) |
| 55-64 | Control | 80 | 50 (62.5) | 27 (33.8) | 3 (3.8) |
| | Messaged | 940 | 565 (60.1) | 342 (36.4) | 33 (3.5) |
| | Total | 1020 | 615 (60.3) | 369 (36.2) | 36 (3.5) |
| 65-74 | Control | 91 | 52 (57.1) | 35 (38.5) | 4 (4.4) |
| | Messaged | 959 | 524 (54.6) | 409 (42.6) | 26 (2.7) |
| | Total | 1050 | 576 (54.9) | 444 (42.3) | 30 (2.9) |
| 75+ | Control | 81 | 42 (51.9) | 37 (45.7) | 2 (2.5) |
| | Messaged | 860 | 403 (46.9) | 439 (51.0) | 18 (2.1) |
| | Total | 941 | 445 (47.3) | 476 (50.6) | 20 (2.1) |

**Supplementary Table 2. Population-level distribution of HbA1c testing status and glycemic categories over 24 weeks.** Percentages are presented with 95% CIs calculated using the Wilson score method.

| | Control (n=5,748) | | Messaged (n=55,494) | |
|---|---|---|---|---|
| **Characteristic** | **Proportion (95% CI), %** | **n** | **Proportion= (95% CI), %** | **n** |
| Not Tested | 92.17 (91.45, 92.84) | 5,298 | 90.86 (90.61, 91.09) | 50,420 |
| Normoglycemia (HbA1c <5.7%) | 5.06 (4.53, 5.66) | 291 | 5.88 (5.69, 6.08) | 3,262 |
| Prediabetes (HbA1c 5.7%-6.4%) | 2.57 (2.20, 3.02) | 148 | 3.07 (2.93, 3.22) | 1,705 |
| Diabetes (HbA1c ≥ 6.5%) | 0.19 (0.11, 0.34) | 11 | 0.19 (0.16, 0.23) | 107 |

**Supplementary Figure 1. Patients randomized in the messaged group received an automated text message encouraging diabetes screening with a link to the study website.**

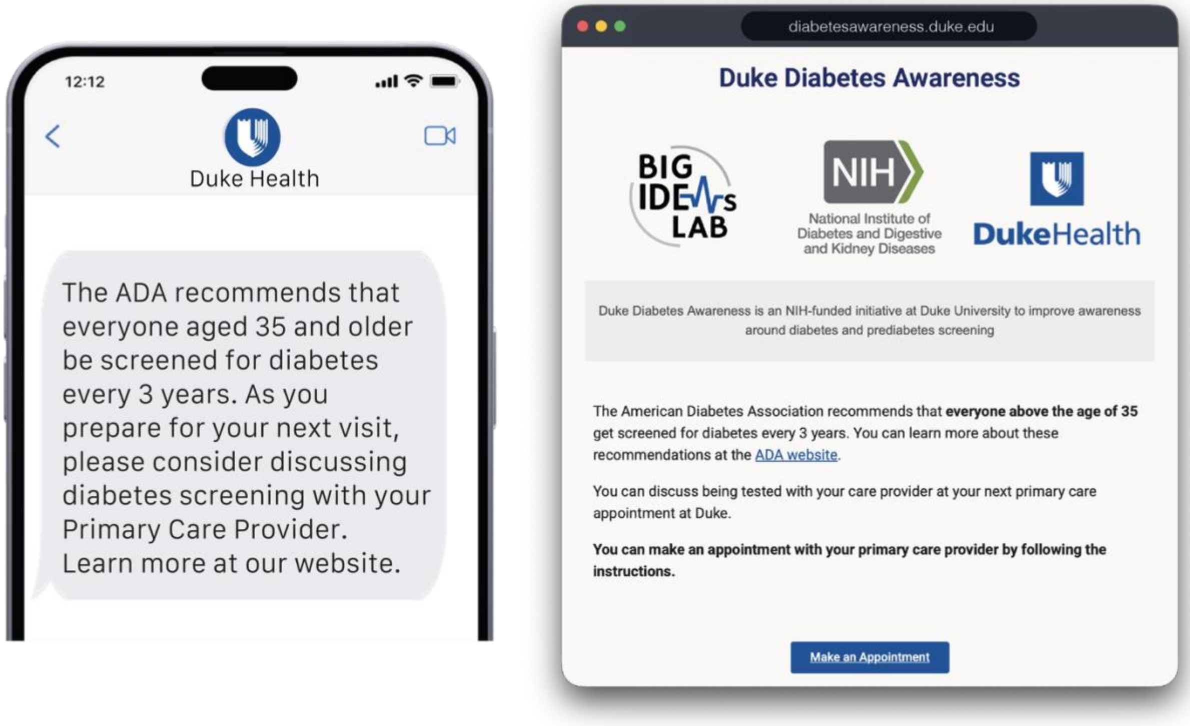

**Supplementary Figure 2. Hazard ratios (HRs) and 95% confidence intervals (CIs) for the primary outcome at 24 weeks, stratified by sex, race, ethnicity, and age group.**

| Subgroup | HR (95% CI) |
|---|---|
| **Biological Sex** | |
| Female | 1.26 (1.12–1.42) |
| Male | 1.09 (0.92–1.28) |
| **Race** | |
| Asian | 1.16 (0.81–1.67) |
| Black or African American | 1.04 (0.81–1.34) |
| Other | 1.07 (0.59–1.94) |
| White | 1.24 (1.10–1.39) |
| **Ethnicity** | |
| Hispanic | 1.76 (0.93–3.32) |
| Not Hispanic/Latino | 1.17 (1.06–1.29) |
| **Age Group** | |
| 35-44 | 1.12 (0.93–1.36) |
| 45-54 | 1.21 (0.96–1.52) |
| 55-64 | 1.21 (0.96–1.52) |
| 65-74 | 1.19 (0.96–1.48) |
| 75+ | 1.23 (0.98–1.54) |

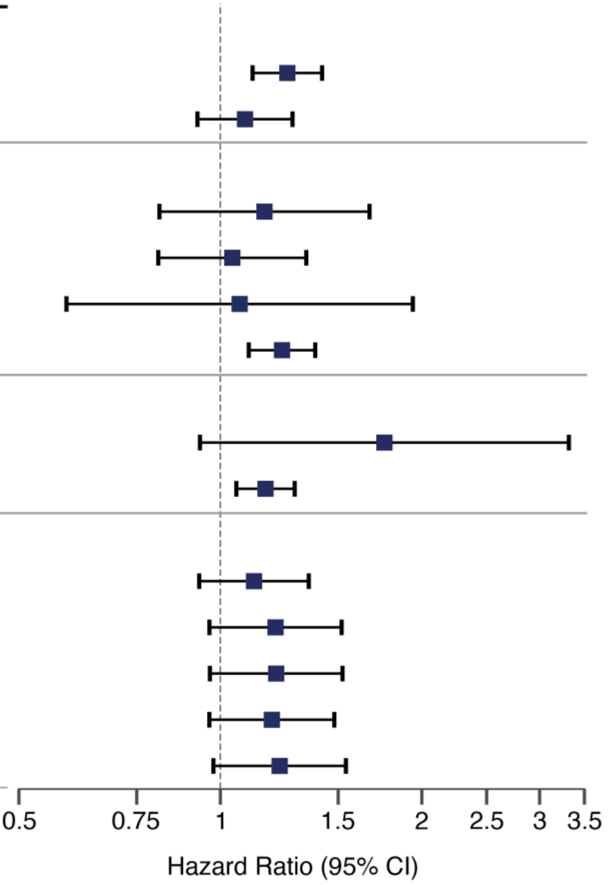
